# Late-life onset psychotic symptoms and incident cognitive impairment in people without dementia: modification by genetic risk for Alzheimer’s disease

**DOI:** 10.1101/2022.09.29.22280446

**Authors:** Byron Creese, Ryan Arathimos, Dag Aarsland, Clive Ballard, Anne Corbett, Zahinoor Ismail

## Abstract

**INTRODUCTION:** Late-life onset psychosis is associated with faster progression to dementia in cognitively normal people, but little is known about its relationship to cognitive impairment in advance of dementia.

**METHODS:** Clinical and genetic data from 2,750 people over 50 without dementia were analyzed. Incident cognitive impairment was operationalized using the IQCODE and psychosis (MBI-psychosis) was rated using the Mild Behavioral Impairment Checklist. The whole sample was analyzed before stratification on APOE-ε4 status.

**RESULTS:** In Cox proportional hazards models, MBI-psychosis had a higher hazard rate (HR) for cognitive impairment relative to the No Psychosis group (HR:3.6, 95% CI:2.2-6, p<0.0001). The HR for MBI-psychosis was higher in APOE-ε4 carriers and there was an interaction between the two (HR for interaction: 3.4, 95% CI:1.2-9.8, p=0.02).

**DISCUSSION:** Psychosis assessment in the MBI framework is associated with incident cognitive impairment in advance of dementia, these symptoms may be particularly important in the context of APOE genotype.

## 1. BACKGROUND

Broadly comprising of visual hallucinations, paranoid delusions, and misidentifications, psychosis occurs in around 40% of people with Alzheimer’s Disease dementia and most people with dementia with Lewy bodies and Parkinson’s disease dementia[1]. While much of the research literature in cognitive aging has focused on symptoms that occur in the context of established dementia, psychotic symptoms do occur in later life in people without dementia. Recognizing the significance of such symptoms, the International Psychogeriatrics Association (IPA) recently revised their criteria for psychosis in neurocognitive disorders to include symptoms occurring in mild neurocognitive disorders (i.e., in advance of dementia in the mild cognitive impairment stage)[2]. Moreover, new research criteria for psychosis in AD from the ISTAART Neuropsychiatric Syndrome Professional Interest Area stipulate that psychosis due to underlying neurodegenerative disease can occur even earlier, in normal cognition[3]. These criteria build upon the development of the Mild Behavioral Impairment (MBI) framework which captures the full spectrum of neuropsychiatric symptoms including apathy, mood/anxiety, impulse dyscontrol, social inappropriateness, and psychosis[4,5]. As a broad syndrome, MBI is linked to cognitive decline and progression to dementia[6–9].

In MCI delusions occur in less than 10% of people while hallucinations are present in 1-2%. In cognitively normal older individuals there is evidence that up to 5-10% of adults over 50 years old have psychotic experiences when assessed in the MBI framework (henceforth MBI-psychosis)[10]. In people with normal cognition, most data are focused on clinically defined groups of patients with psychotic disorders. Overall, these tend to show an increased risk of dementia, however data on individuals whose symptoms are not attributable to another clinical disorder are scarce[11–14]. What data there are, are consistent with the above and suggest that psychosis confers the highest risk for dementia of all neuropsychiatric syndromes in cognitively normal people[15,16].

With an intensifying need to identify early markers of neurodegenerative disease it is imperative that further characterization of a low frequency but seemingly high-risk syndrome like psychosis is carried out in cognitively normal samples. Key avenues relate to the impact of psychosis on functional outcomes and identifying the most clinically relevant groupings. Thus, in the present study we examined 1) the relationship between MBI-psychosis and incident cognitive impairment operationalized by the Informant Questionnaire on Cognitive Decline in the Elderly (IQCODE); 2) whether this relationship was modified by genetic risk for AD and sex. This analysis complements emerging computerized testing data as the IQCODE has a reference period of 10 years, is not confounded by education or intelligence, and focuses on cognitive impairments that can give a better picture of the impact of symptoms on everyday functioning.

## 2. METHODS

### 2.1. Data source and participant selection

Data used in this analysis are from PROTECT (www.protectstudy.org.uk; Research Ethics Committee reference number 13/LO/1578), an online study that aims to examine lifestyle, mental health, and genetic risk factors for cognitive aging. It was launched in 2015 and the data analyzed here are taken from a January 2022 data freeze. Written informed consent from participants is obtained online. Participants may nominate a study partner who also answers questionnaires relating to the participant. All assessments are completed online and annually (within a 6-month window of the anniversary date). On enrolling to the study, participants are asked to confirm that they do not have a diagnosis of dementia have access to a computer and internet, are aged 50 years or over, and able to read and write English.

All participants who nominated a study partner, had available genetic data and were cognitively normal were considered for analysis. Normal cognition was defined as having baseline IQCODE<3.6 (see below for more detail) and answering ‘no’ to the question “Have you ever received a diagnosis from a medical professional of mild cognitive impairment (MCI)?”). Only individuals who enrolled on or before 1^st^ February 2017 were included in the analysis. This was to ensure that per protocol follow up time was at least 5 years; an appropriate time for a cognitive normal, relatively young sample. Further exclusions were applied as detailed below and in the CONSORT diagram in Figure 1.

**Figure 1:**
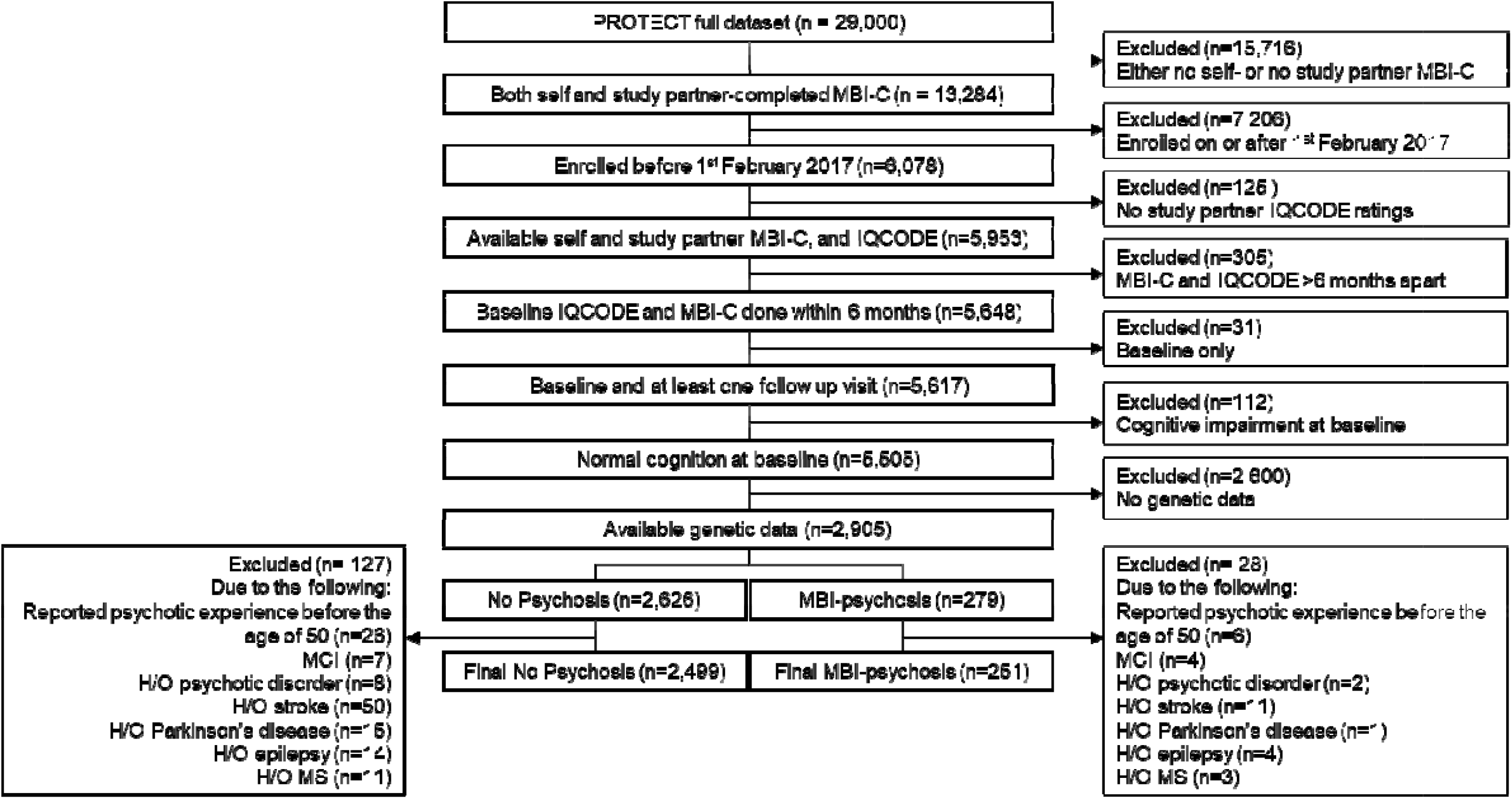
CONSORT chart showing participant selection.

### 2.2. Demographic and clinical history

Age, gender, education level, ethnicity and, the presence of any hearing or vision problems (“Do you have problems with your hearing?” and “Do you have any visual impairment that requires you to wear glasses/contact lenses?”) at baseline were recorded. Self-reported history of the following conditions was also obtained by asking if the participant had ever received a diagnosis of: hypertension, stroke, heart disease/attack/angina, diabetes, MCI, Parkinson’s disease, high cholesterol, hypothyroidism, hyperthyroidism, arthritis, Huntington’s disease, current cancer, remitted cancer, osteoporosis, asthma, epilepsy, motor neuron disease, multiple sclerosis, Paget’s disease, deep vein thrombosis, HIV/AIDS, hepatitis C. Similarly, self-reported history of the following psychiatric conditions was also recorded: depression, mania/bipolar depression, anxiety/generalized anxiety disorder, social anxiety disorder, agoraphobia, panic attacks, obsessive compulsive disorder, anorexia nervosa, bulimia nervosa, binge eating, schizophrenia, any other type of psychotic illness, personality disorder, autism spectrum disorder, attention deficit and hyperactivity disorder. The presence of schizophrenia or any other psychotic disorder was coded separately while all other mental health conditions were coded collectively as ‘history of mental health condition.’ Finally, four questions were asked that pertain to the presence of psychotic experiences during the participant’s lifetime. These cover hallucinations (visual and auditory) and delusions (the full questions are listed in supplementary material). Following these participants were also asked at what age they first had such experiences. These questions are asked separately to the MBI-C and provide an additional screening measure to rule out people with early life psychotic experiences.

### 2.3. Primary outcome: incident cognitive impairment

Because this study is concerned with cognitive impairment in advance of dementia, we used the study partner-rated IQCODE scale. The IQCODE is a 16-item scale where respondents are asked to rate change in cognitive ability on a scale of 1 (much better) to 5 (much worse) across a range of everyday activities, with 3 representing no change. The full list of questions is included in the supplement. The final IQCODE score is obtained by taking the mean of the 16 items. Various cut points are used to capture cognitive impairment, typically ranging from 3.3 to 3.6. For this study, IQCODE >3.3 and >3.6 were considered[17]. To inform choice of cognitive endpoint we tested the association of each with APOE-ε4 status in the whole PROTECT sample. In logistic regression analysis with each IQCODE cut point as the binary dependent variable, carrying two copies of the APOE-ε4 allele was associated with worse cognition relative to no copies but there was a modestly higher odds ratio (OR) associated with a cut point of 3.6 so this was chosen as the primary end point (>3.6: OR=2.21, 95% CI:1.22-3.71, p=0.005; >3.3: OR=1.6, 95% CI:1.12-2.24, p=0.008). For context, a mean score of >3.6 can be reached by rating ‘much worse’ (i.e., 5) on at least 5/16 items or ‘a bit worse’ (i.e., 4) on 10/16 items (assuming all others are rated as ‘no change’). Follow up times were calculated from baseline to date of outcome, censoring, or 5 years for those who remained cognitive unimpaired throughout.

### 2.4. MBI-psychosis

Psychotic symptom status was ascertained from the Mild Behavioral Impairment Checklist (MBI-C)[18]. Ratings were obtained from both participants and study partners. A total of 33 questions capture five domains (mood, apathy, impulse dyscontrol, social inappropriateness, and psychosis). There are five questions pertaining to psychosis, three covering delusion-type experiences (paranoid, harm, and grandiose-type) and two for hallucinations (visual and auditory).

Based on the ratings of participants and study partners (which had to be within six months of each other, see Figure 1), two groups were created: MBI-psychosis and No Psychosis. Participants were classified as MBI-psychosis if they or their study partner rated any of the five psychosis items as present at their first visit. Participants were coded as No Psychosis if they scored zero on all five items on both participant and study partner ratings. To reflect MBI diagnostic criteria (which stipulate symptoms should not be attributable to another medical condition) participants with Parkinson’s disease, epilepsy, multiple sclerosis or history of stroke were excluded (see Figure 1). Moreover, although the MBI-C instructions state that symptoms should be new onset, there were 33 participants who reported having a psychotic experience under the age of 50, these were excluded to ensure only late-life psychosis cases in our sample. MBI-C ratings were completed within six months of the initial IQCODE assessment.

### 2.5. Genotype quality control, APOE determination and polygenic risk score calculation

A detailed description of genotyping, quality control (QC) is provided in the supplement and described previously[19]. Genotype data were available for 9,146 PROTECT study participants. Individual-level QC steps included call-rate (98%) filtering, relatedness, excess heterozygosity and gender mismatch. Individuals not of European ancestry were excluded. These steps led to the removal of 790 people. Variant-level QC included call-rate (98%), Hardy-Weinberg deviation (p<0.00001). Genotypes were imputed to 1000 Genomes European reference panel using the Michigan imputation server and genotype phasing using Eagle. Variants were restricted to single nucleotide polymorphisms (SNPs) only, with a MAF > 0.001. An absolute cut-off of 0.7 was applied to the imputation quality of variants (*R*^*2*^ as reported by the Michigan imputation server). The number of variants remaining after quality control was 9,415,055.

In addition to APOE genotype, a polygenic risk score (PRS) for AD was calculated which also captures non-APOE genetic risk. AD PRS was calculated using International Genomics of Alzheimer’s Project (IGAP) AD genome-wide association study (GWAS) summary statistics using PRSice v2.2.12 (broadly, this is generated by calculating the effect size-weighted sum of the number of risk alleles carried at each SNP by an individual)[20]. All available SNPs from the IGAP study were used (83,540) to calculate the PRS, this score was associated with cognitive performance on computerized testing in a sample that overlaps with the current one[21]. AD PRS tertiles were generated in the whole sample before exclusions.

### 2.6. Statistical analysis

All analyses were carried out in R version 4.1.3. Baseline demographic, clinical and genetic variables were compared between the MBI-psychosis and No Psychosis groups using t-tests (or Mann-Whitney test for non-parametric data) or Fisher’s exact test.

Survival to cognitive impairment was compared using Kaplan-Meier (KM) survival curves and log-rank test over a maximum follow up period of 5 years. Following this, Cox proportional hazards models were fitted to the data and Hazard Ratios (HR) were generated to describe difference in rates of cognitive impairment in the MBI-psychosis group relative to the No Symptom group. Additional covariates were gender, age, history of any mental health condition and APOE-ε4 status.

The sample was then stratified on APOE-ε4 status, and then separately on gender (informed by previous research showing interactions between gender and psychosis on cognitive decline[22]), and survival for MBI-psychosis groups explored in each strata. Finally, interaction terms (MBI-psychosis*APOE-ε4 then MBI-psychosis*gender) were fitted. To explore the role of non-APOE genetic variation, HRs for MBI status were generated in each AD PRS tertile. Covariates remained as above but to evaluate the contribution of other non-psychosis MBI items, MBI-C total score excluding the five psychosis items was entered as a covariate in a second adjusted model.

Analysis was completed using the *survminer* package for KM plots and the *survival* package for Cox proportional hazards models. Proportional hazards assumptions were tested and confirmed using the *cox*.*zph* function.

## 3. RESULTS

### 3.1. Participant demographics and clinical characteristics

After the exclusions detailed in Figure 1, a total of 2,750 participants were included in the final analytical sample (mean age 64±6.8; 2,165 - 74% - women). There were 2,499 participants classified as having No Psychosis and 251 in the MBI-psychosis group (Table 1). The MBI-psychosis and No Psychosis groups did not statistically differ on any demographic variables. There was, however, a numerically higher proportion of individuals with undergraduate degrees in the No Psychosis group (37% vs. 29%). In terms of self-reported (note this may therefore differ from genetically determined ancestry) ethnicity, virtually all participants reported being white (either British - including English, Scottish and Welsh, Irish, other European or non-European) with five reporting being of mixed-race background.

**Table 1:** Baseline sample characteristics by MBI-psychosis status.

|  | No Psychosis |  | MBI Psychosis |  |
| --- | --- | --- | --- | --- |
| **n** | 2499 |  | 251 |  |
|  | Mean | SD | Mean | SD |
| **Age** | 64 | 6.8 | 63 | 7.0 |
| **Gender** | n | % | n | % |
| Male | 650 | 26 | 66 | 26 |
| Female | 1849 | 74 | 185 | 74 |
|  | Median | IQR | Median | IQR |
| **Proxy-rated MBI-C total score excluding psychosis items (median, IQR)** | 1 | 4 | 6 | 9 |
| **Self-rated MBI-C total score excluding psychosis items (median, IQR)** | 0 | 2 | 3 | 9 |
|  | n | % | n | % |
| **Delusions<sup>1</sup>** | - | - | 234 | 93 |
| **Hallucinations<sup>1</sup>** | - | - | 20 | 8 |
|  | n | % | n | % |
| **Number of APOE-ε4 alleles** |  |  |  |  |
| 0 | 1733 | 69 | 160 | 64 |
| 1 | 702 | 28 | 85 | 34 |
| 2 | 64 | 3 | 6 | 2 |
|  | n | % | n | % |
| **Education** |  |  |  |  |
| Left school at 16 | 290 | 12 | 39 | 16 |
| Left school at 18 | 290 | 12 | 29 | 12 |
| Vocational qualification | 467 | 19 | 54 | 22 |
| Undergraduate degree | 923 | 37 | 72 | 29 |
| Post-graduate or doctoral degree | 529 | 21 | 57 | 23 |
|  | n | % | n | % |
| **Ethnicity<sup>2</sup>** |  |  |  |  |
| White | 2494 | 100 | 251 | 100 |
| Mixed race (White and Black African) | 2 | 0 | 0 | 0 |
| Any other mixed ethnicity<sup>3</sup> | 3 | 0 | 0 | 0 |
|  | n | % | n | % |
| **Medical history<sup>5</sup>** |  |  |  |  |
| Wears glasses or contact lenses | 2,390 | 96 | 232 | 92 |
| Hearing problem | 723 | 29 | 77 | 31 |
| Any mental health condition<sup>6</sup> | 873 | 35 | 115 | 46 |
| Hypertension | 696 | 28 | 81 | 32 |
| Heart disease | 150 | 6 | 20 | 8 |
| Diabetes | 95 | 4 | 12 | 5 |
| High cholesterol | 657 | 26 | 64 | 25 |
| Hypothyroidism | 176 | 7 | 21 | 8 |
| Hyperthyroidism | 50 | 2 | 3 | 1 |
| Arthritis | 523 | 21 | 59 | 24 |
| Cancer (current) | 50 | 2 | 4 | 2 |
| Cancer (full remission) | 207 | 8 | 29 | 12 |
| Osteoporosis | 154 | 6 | 17 | 7 |
| Asthma | 207 | 8 | 27 | 11 |
| Paget's disease | 1 | 0 | 0 | 0 |
| Deep vein thrombosis | 32 | 1 | 2 | 1 |
| Hepatitis C | 2 | 0 | 0 | 0 |

Abbreviations: MBI: Mild Behavioral Impairment; MBI-C: Mild Behavioral Impairment Checklist.
<sup>1</sup>Does not sum to 235 because 3 participants had both delusions and hallucinations.
<sup>2</sup>Refers to self-reported ethnicity not genetically determined ancestry. Only non-zero frequency items are presented but the following options were available: white: English / Welsh / Scottish / Northern Irish / British; White: Irish; White: Gypsy or Irish Traveler; White: European; White: Non-European; Mixed: White and Black Caribbean; Mixed: White and Black African; Mixed: White and Asian; Mixed: Any other Mixed / Multiple ethnic background; Asian / Asian British: Indian; Asian / Asian British: Pakistani; Asian / Asian British: Bangladeshi; Asian / Asian British: Chinese; Asian / Asian British: Any other Asian background; Black / African / Caribbean / Black British: African; Black / African / Caribbean / Black British: Caribbean; Any other Black / African / Caribbean background; Other ethnic group: Arab; Any other ethnic group.
<sup>3</sup>White includes any of the 'white' options listed in footnote 1.
<sup>4</sup>Denotes any other mixed ethnic background besides those listed in footnote 1.
<sup>5</sup>Only conditions rated as present by at least one person are shown, see text for full list of conditions screened for.
<sup>6</sup>Excludes schizophrenia and history of any other psychotic illness.

The median proxy-rated MBI-C total score excluding psychosis items was 6 for MBI-psychosis and 1 for No Psychosis (W=131215, p<0.0001). The median self-rated MBI-C score excluding psychosis items was 3 for MBI-psychosis and 0 No Psychosis (W=201904, p<0.0001). Ninety-three percent (234) of MBI-psychosis participants had delusions while 8% (20) had hallucinations. Only three participants experienced both symptoms. There was a statistical difference between the two groups on history of any mental health condition (MBI-psychosis 46% vs. No Psychosis 35%, Fisher’s exact p=0.0008; note this does not include schizophrenia and other psychotic disorders as these individuals were excluded from the analysis). All other self-reported medical conditions were comparable between the two groups (Table 1).

### 3.2. Survival analysis

In the whole sample, while cognitive impairment was an uncommon event, the MBI-psychosis group had significantly lower survival than the No Psychosis group (five-year survival probability for the MBI-psychosis group: 86% [95% CI:79%-94%]; for No Psychosis: 94% [95% CI:91%-96%]; log-rank p<0.0001).

The sample was then stratified by APOE status. Due to the very small numbers of ε-4 homozygotes, two strata were created: no ε-4 alleles (non-carrier, n=1,893) and one or two ε-4 alleles (ε-4 carrier, n=857). Figures 2 A and B show the KM curve for cognitive impairment free survival probability over 5 years for MBI-psychosis compared with No Psychosis in APOE-ε4 non-carriers (A) and carriers (B). In APOE-ε4 carriers, MBI-psychosis was associated with a lower survival probability relative to the No Psychosis group (81% [95% CI:71%-92%] vs. 94% [95% CI:90%-98%, log rank p<0.0001). There was no difference in survival probability between the two groups in APOE-ε4 non-carriers (89% [95% CI:80%-99%] vs. 94% [93% CI:91%-96%, log rank p=0.06).

**Figure 2:**
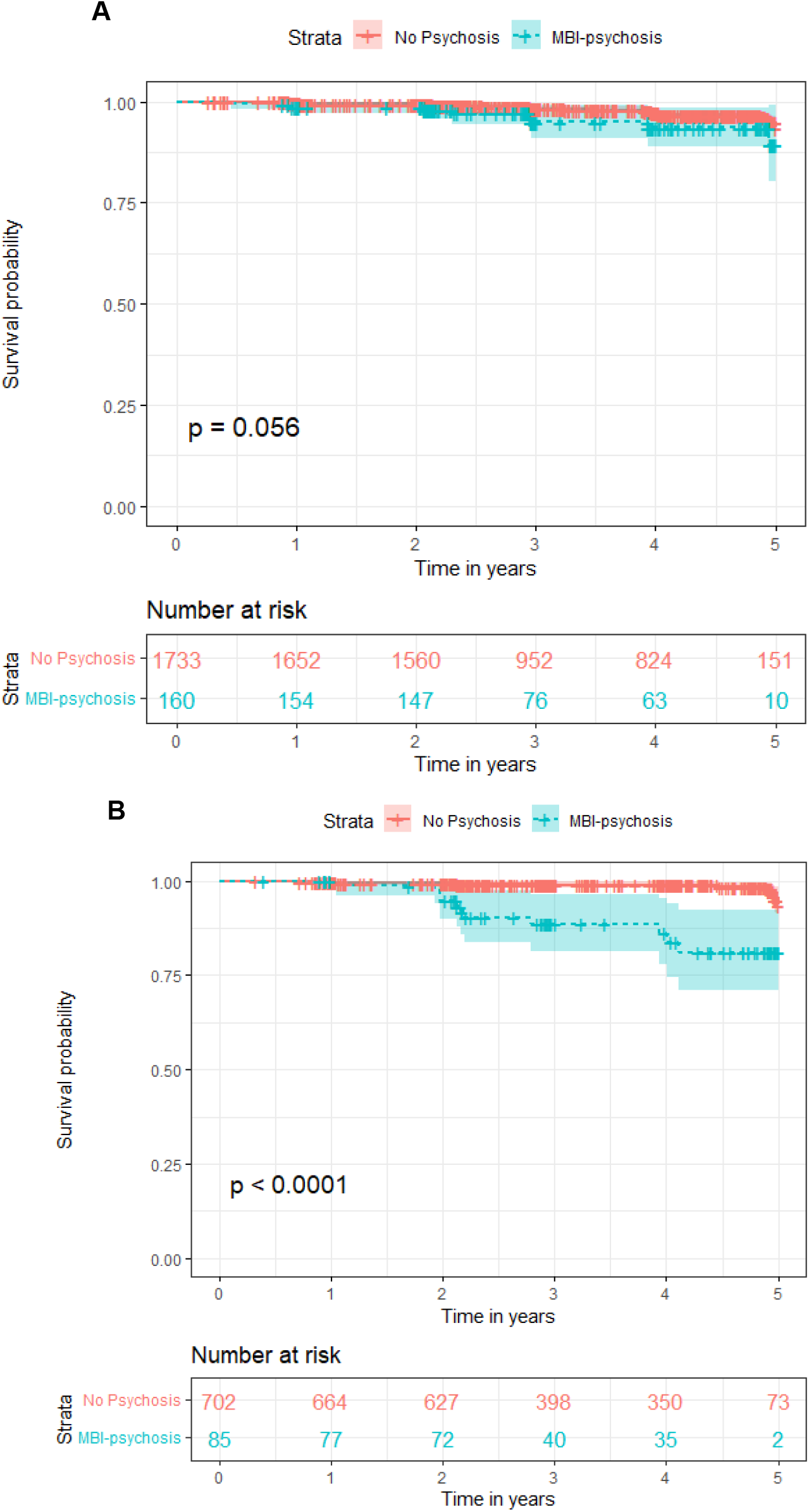
**KM curve for the probability of remaining cognitive impairment free over five years, stratified by MBI-psychosis status in APOE-ε4 noncarriers (A) and carriers (B). P-values are from the log-rank test.**

### 3.3. Cox proportional hazards analysis

Full lists of regression coefficients, standard errors and p-values are included in the supplement.

#### 3.3.1. Whole sample

The MBI-psychosis group had a 3.6-fold higher hazard ratio than the No Psychosis group (HR:3.6, 95% CI:2.2-6, p<0.0001). Eight percent (n=20) of the MBI-psychosis group progressed to cognitive impairment, compared with 2.6% (n=65) of the No Psychosis group (see Supplementary Figure 1 for forest plot of adjust HRs).

#### 3.3.2. Stratified analyses

Figure 3 and Supplementary Figure 2 show the whole sample stratified by APOE status and gender respectively.

**Figure 3:**
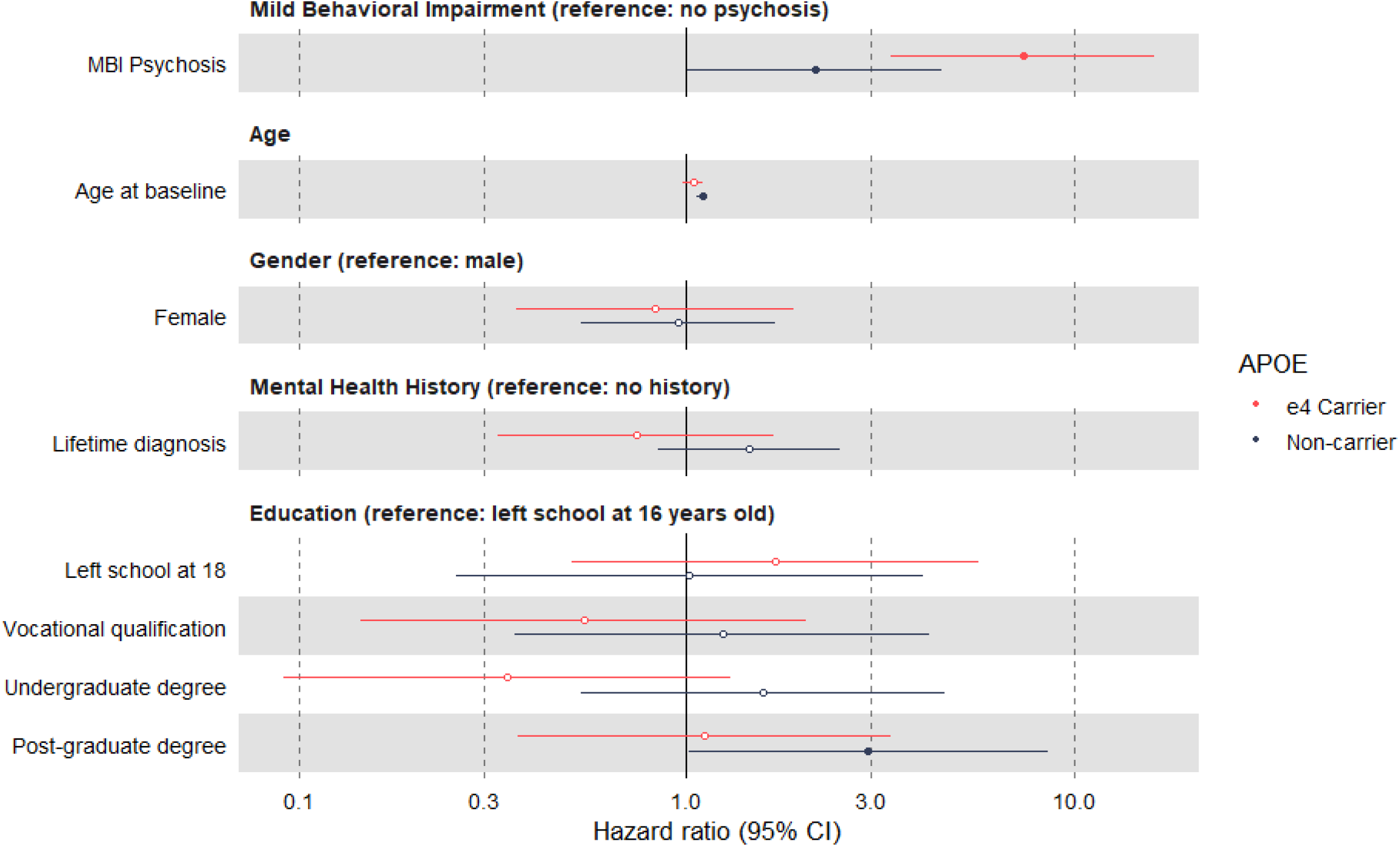
**Forest plot of adjusted hazard ratios for cognitive impairment across all covariates, stratified by APOE carrier status (0 vs. 1 or 2 alleles). Filled points denote statistical significance at p<0.05.**

##### 3.3.2.1. Genetic risk for AD

###### APOE

There was a 2-fold higher hazard for cognitive impairment in the MBI-psychosis group relative to the No Psychosis group in people with no APOE ε-4 alleles but this did not reach statistical significance (HR:2.1, 95% CI:1-4.4, p=0.05). In the ε-4 carrier group, the MBI-psychosis group had a 7.3-fold greater hazard rate than the No Psychosis group (HR:7.3, 95% CI:3.4-16.2, p<0.0001). The interaction between MBI-psychosis group and APOE carrier status was statistically significant (HR:3.4, 95% CI:1.2-9.8, p=0.02, Figure 3).

###### AD PRS

The hazard for cognitive impairment associated with MBI-psychosis relative to No Psychosis was comparable across the three PRS tertiles. In the lowest tertile, there was a 3-fold higher hazard rate for cognitive impairment in the MBI-psychosis group relative to the No Psychosis group (HR:3.4, 95% CI:1.3-8.5, p=0.01). In the middle tertile, there was a 4.4-fold higher hazard rate in the middle tertile (HR:4.4, 95% CI:2-9.6, p=0.0002). In the top tertile there was a 3-fold higher hazard rate in the top tertile (HR:3, 95% CI:1.1-8.2, p=0.02). There was no interaction between MBI-psychosis status and AD PRS tertile.

A cross-tabulation of APOE carrier status by AD PRS tertile shows that a significant proportion of ε-4 carriers are present across the lowest, middle, and highest tertiles (24%, 33% and 37% respectively). Thus, the APOE signal is spread across the total PRS strata which may explain the difference in results between the APOE strata and the PRS strata. To explore this further post-hoc, we examined a second AD PRS that only included those SNPs showing genome-wide significant associations with AD (i.e. 22 SNPs, including the APOE locus and calculated as previously described [19]). As would be expected, in the top tertile of this PRS, 76% of people carried at least one APOE-ε4 allele compared to 13% of the middle tertile and 1% of the bottom tertile. Here, we observed a similar interaction as for APOE (MBI-psychosis*top tertile: HR:7, 95% CI:1.4-3.5, p=0.02; MBI-psychosis*middle tertile: HR:3.4, 95% CI:0.6-1.9, p=0.6).

When controlling for other neuropsychiatric symptoms, MBI-psychosis was no longer associated with a higher hazard rate for cognitive impairment in APOE ε-4 non-carriers (HR:0.9, 95% CI:0.4-2, p=0.8). In APOE ε-4 carriers the HR was reduced from 7.4 to 3.6 (95% CI:1.6-8.4). The interaction between MBI-psychosis group and APOE carrier status remained statistically significant (HR:4.2, 95% CI:1.4-12.1, p=0.009).

##### 3.3.2.2. Gender

The hazard rate for cognitive impairment was 5.2-fold (95% CI:2.2-12.4, p=0.0001) higher in the MBI-psychosis group relative to the No Psychosis group in men and 2.9-fold (95% CI:1.5-5.6, p=0.001) higher than No Psychosis in women but the interaction did not reach statistical significance (HR:0.6, 95% CI:0.2-1.6, p=0.3, Supplementary Figure 2).

## 4. DISCUSSION

These data add to converging evidence that highlights the importance of late-life onset psychotic symptoms in cognitive aging. Although uncommon, occurring in less than 10% of the sample (with only 0.7% experiencing hallucinations), psychotic symptoms in cognitively normal individuals were found to have a 3.6-fold higher risk of incident cognitive impairment. By using the IQCODE as an outcome, this study confirms is the impact of such symptoms on cognitive abilities that affect daily life in advance of dementia, building upon studies that have examined cognitive trajectories or those that have followed individuals to the point of clinical dementia diagnosis[15,16,22].

We found evidence of effect modification by APOE status; a significantly greater HR for incident cognitive decline in the MBI-psychosis group was present only in individuals who carry at least one ε-4 allele. This relationship was not wholly attributable to other MBI symptoms. APOE is a well-established risk factor for AD and has been show to interact with affective neuropsychiatric symptoms have to influence dementia risk, though ours is the first study to show an interaction with psychosis on cognitive impairment in advance of dementia[23]. These findings bring a new perspective to the broader relationship between APOE and cognitive decline by suggesting that the presence of psychotic symptoms among carriers may be of particular clinical interest.

It should be highlighted that we did not observe an interaction when analyzing AD PRS (which includes APOE genotype) by tertile. It is important to note that 24% of the lowest tertile were APOE carriers, a substantial proportion even relative to the middle and high tertiles (33% and 37% respectively). A possible explanation for this discrepancy is that the effect observed in the main analysis is APOE-specific, and thus present to a degree across all three PRS tertiles. In addition to the two APOE SNPs, there are >83,500 other SNPs which were used to determine each person’s AD PRS (and by extension their genetic risk tertile) and it appears that these are not clearly associated with the cognitive impairment outcome used in this study. This is backed up by our post-hoc analysis of a 22 SNP PRS (i.e., with far fewer non-APOE SNPs to influence the score) where we observe an interaction similar to that shown in the main APOE analysis. This is in line with a number of other studies which clearly show variation in findings according to which measure of genetic risk is used (a PRS with just a few SNPs, tens of thousands of SNPs, or APOE alone)[19,24–28].

There was no effect modification by gender in this sample. In a recent study of cognitive trajectories on computerized testing in a sample that overlaps with this one, the association between cognitive decline and psychosis was only present in men. It is possible that moderation by gender does not extend to the IQCODE, which reflects cognitive function assessed by decline in daily activities rather than precise measurement of specific cognitive domains. Alternatively, the nominal difference in this sample may be due to the low frequency of the cognitive outcome, and thus the lack of precision of the estimate.

To our knowledge this is the first study to evaluate the impact psychotic symptoms assessed specifically in the MBI framework using the MBI-C, drawing on both participant and study partner ratings to capture symptoms fully. This has allowed us to assess new onset and persistent symptoms more precisely and in accordance with MBI criteria. The online nature of this study represents a key strength, allowing us to reach participants with MBI who would most likely not be in contact with clinical services however we acknowledge with this comes limitations. Among these, are the overrepresentation of women, those reporting White ethnicity and those with a higher education. These are common limitations to large studies which rely on self-selection, and this underscores the importance of data collection in more diverse samples. These data should motivate such studies.

In summary, by reporting an increased risk of cognitive impairment and an interaction with genetic risk for AD, this study underscores the importance of psychotic symptoms assessed in the Mild Behavioral Impairment framework - i.e., sustained, later-life onset and not attributable to other psychiatric conditions - in cognitive aging.

## Data Availability

All data produced in the present study are available via request to the PROTECT study.

## Conflicts of Interest

Clive Ballard has received contract grant funding from ACADIA, Lundbeck, Takeda, and Axovant pharmaceutical companies and honoraria from Lundbeck, Lilly, Otsuka, and Orion pharmaceutical companies. Dag Aarsland has received research support and/or honoraria from Astra-Zeneca, H. Lundbeck, Novartis Pharmaceuticals, and GE Health, and serves as a paid consultant for H. Lundbeck and Axovant. Zahinoor Ismail has received honoraria/consulting fees from Janssen, Lundbeck, and Otsuka, although not related to this work.

## Acknowledgements

This research was also supported by the NIHR Collaboration for Leadership in Applied Health Research and Care South West Peninsula. The views expressed are those of the author(s) and not necessarily those of the NHS, the NIHR or the Department of Health and Social Care. Genotyping was performed at deCODE Genetics. The authors would like to acknowledge the use of the research computing facility at King’s College London, Rosalind (https://rosalind.kcl.ac.uk), which is delivered in partnership with the NIHR Biomedical Research Centres at South London & Maudsley and Guy’s & St. Thomas’ NHS Foundation Trusts, and part-funded by capital equipment grants from the Maudsley Charity (award 980) and Guy’s & St. Thomas’ Charity (TR130505).

## Funding

This work was funded in part through the MRC Proximity to Discovery: Industry Engagement Fund (External Collaboration, Innovation and Entrepreneurism: Translational Medicine in Exeter 2 (EXCITEME2) ref. MC_PC_17189) awarded to Dr Creese. This paper represents independent research part funded by the National Institute for Health Research (NIHR) Biomedical Research Centre at South London and Maudsley NHS Foundation Trust and King’s College London. None of the funding bodies had any role in the design, analysis or interpretation of data, or the drafting of the manuscript.

## Notes

### Author Declarations

London Bridge NHS Research Ethics Committee has approved this research (Ref: 13/LO/1578)

